# Beyond Deaths per Capita: Comparative CoViD-19 Mortality Indicators

**DOI:** 10.1101/2020.04.29.20085506

**Authors:** Patrick Heuveline, Michael Tzen

## Abstract

**Objectives:** Following well-established practices in demography, this article discusses several measures based on the number of CoViD-19 deaths to facilitate comparisons over time and across populations.

**Settings:** National populations in 186 UN countries and territories and populations in first-level sub-national administrative entities in Brazil, China, Italy, Mexico, Peru, Spain, and the USA.

**Participants:** None (death statistics only).

**Primary and Secondary Outcome Measures:** An unstandardized occurrence/exposure rate comparable to the Crude Death Rate; an indirectly age-and-sex standardized rate that can be derived even when the breakdown of CoViD-19 deaths by age and sex required for direct standardization is unavailable; the reduction in life expectancy at birth corresponding to the 2020 number of CoViD-19 deaths.

**Results:** To date, the highest unstandardized rate has been in New York, at its peak exceeding the state 2017 Crude Death Rate. Populations compare differently after standardization: while parts of Italy, Spain and the USA have the highest unstandardized rates, parts of Mexico and Peru have the highest standardized rates. For several populations with the necessary data by age and sex for direct standardization, we show that direct and indirect standardization yield similar results. US life expectancy is estimated to have declined this year by more than a year (−1.26 years), far more than during the worst year of the HIV epidemic, or the worst three years of the opioid crisis, and to reach its lowest level since 2008. Substantially larger reductions, exceeding two years, are estimated for Panama, Peru, and parts of Italy, Spain, the USA, and especially, Mexico.

**Conclusions:** With lesser demand on data than direct standardization, indirect standardization is a valid alternative to adjust international comparisons for differences in population distribution by sex and age-groups. A number of populations have experienced reductions in 2020 life expectancies that are substantial by recent historical standards.

**Strengths and limitations:**

- The CoViD-19 mortality indicators presented in this article are directly comparable with three well-established indicators of overall mortality: the Crude Death Rate, the Age-Standardized Death Rate and Life Expectancy at Birth
- In particular, this article demonstrates that when CoViD-19 deaths in a population are not tabulated by sex and age, indirect standardization techniques can still be used to improve comparisons of CoViD-19 mortality in this and other populations by accounting for differences in population distributions by age and sex
- While requiring additional data on mortality from other causes, translating cumulative numbers of CoViD-19 deaths into their impact on life expectancy at birth allows for comparison of CoViD-19 mortality with previous reversals in secular mortality declines
- The comparability of these CoViD-19 mortality indicators is affected by potential differences in identifying and reporting CoViD-19 as a cause of death across populations
- Further analyses are needed to assess potential changes in mortality from other causes induced by CoViD-19, as those would also contribute to the impact of CoViD-19 on life expectancy at birth

## Background

As of June 1^st^, deaths from the novel coronavirus disease 2019 (CoViD-19) had been reported in 186 of the 235 countries and territories of the United Nations system (UN). As with previous pandemics,^1^ the disease progression can be more reliably tracked with death than with case counts. Cumulative CoViD-19 death counts at a given time depend on the determination of the cause of death, delays in reporting deaths to central reporting agencies—different for deaths at home, in hospitals and other institutions—and delays in verification, consolidation and publication at reporting agencies. In the USA, for instance, the grim milestone of 100,000 cumulative CoViD-19 deaths was reached at the end of May, when data from the Center for Disease Control and Prevention (CDC) suggested that the number of deaths in the country exceeded expectations based on past trends by about 130,000.^2^ While CoViD-19 deaths might not be fully reported, the death undercount is both easier to estimate and an order-of-magnitude smaller than the proportion of unreported cases. CDC data from large-scale seroprevalence surveys suggest that as much as 10 times more SARS-CoV-2 infections occurred than the number of reported CoViD-19 cases^3^—a situation in no way unique to the US.^4^ CoViD-19 mortality indicators are also more pertinent for assessing public-health measures that were intended less to reduce the eventual number of cases than to “flatten the curve” and eventually limit the number of CoViD-19 deaths by keeping the need for emergency hospitalizations below local hospital capacity.

For comparative purposes, cumulative death counts are affected by several demographic characteristics such as, most obviously, population size. The deaths per capita ratio, however, represent the first rather than the only adjustment that can be taken towards more meaningful CoViD-19 mortality comparisons. Following well-established practices in demography,^5^ this article presents more refined indicators that can be derived with additional demographic data. The corresponding measures are discussed using results for the 186 UN countries and territories with at least one death by June 1^st^. To illustrate issues of scale, the measures are also calculated at the first sub-national administrative level (e.g., states or provinces) in selected countries, which were the largest countries in the successive “epicenters” of the pandemic over time: first China, then Italy and Spain, followed by the US, and now Brazil, Mexico and Peru. Altogether, at least one of the measures presented here is estimated for a total of 386 national and subnational populations.

## Methods and Data

We first calculate an occurrence/exposure *rate* that relates the cumulative number of CoViD-19 deaths to the number of person-years lived in the population during the period. With the standard approximation for person-years, the period Crude CoViD-19 Death Rate (*CCDR*) can be measured as:

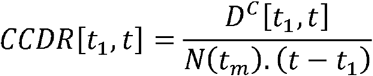

where *t*_1_ is an initial time, *D*^*C*^[*t*_1_,*t*] a cumulative CoViD-19 deaths count between times *t*_1_ and *t*, and *N*(*t*_*m*_) an estimate of the total population size at time *t*_*m*_ between time *t*_*1*_ and time *t*. The difference between this period rate and the deaths per capita ratio is easy to miss when the deaths count in the numerator, identical for both, is an annual number of deaths. In that case, the number of person-years in the denominator of the occurrence/exposure rate can indeed be approximated by the population size at some point during the year. However, the two are no longer directly comparable, and the metric of the ratio difficult to interpret, when the deaths counts correspond to periods of different durations. On the contrary, the *CCDR* is expressed in deaths per person-*year* and remains directly comparable to the annual Crude Death Rate (*CDR*) available for most populations. We first calculate the *CCDR* for the period starting on the day of the first death in the population, which was obtained from World Health Organization (WHO) daily situation reports,^6^ and ending on January 1^st^, 2021. The cumulative number of deaths reported up to that day was obtained from Johns Hopkins University’s Center for Systems Science and Engineering (CSSE)^7^ and total population size was obtained from the UN.^8^ (Additional sources used for sub-national units are referenced in the Technical Appendix.)

As age and sex variations in CoViD-19 mortality have been clearly established,^9^ the period rates should be adjusted to take into account differences in age and sex distributions. Direct age-and-sex standardization requires data on CoViD-19 deaths by age and sex, which are unavailable or unreliable for a majority of UN countries and territories and most sub-national populations. An alternative approach, known as indirect standardization, borrows an age-and-sex pattern of mortality from a well-documented population so that only the age-and-sex distribution of the populations of interest is required. Based on this approach, we calculate the Comparative CoViD-19 Mortality Ratio (*CCMR*):

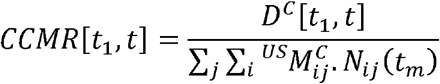

where ^*US*^*M*_*ij*_^*C*^ is the CoViD-19 death rate specific to age group *i* and sex *j* in the US and *N*_*ij*_(*t*_*m*_) is the size of the age group *i* for sex *j* in the population of interest. The reference age-and-sex death rates selected here to illustrate the technique were obtained from the Centers for Disease Control and Prevention (CDC) weekly-updated distribution of CoViD-19 deaths by age and sex in the US,^10^ to date the largest number of CoViD-19 deaths distributed by age and sex. Unavailable only for the 13 countries/territories whose population size is less than 90,000, population age- and-sex distributions were taken from the UN data and, for subnational populations, national statistics.

Multiplying a population *CCMR* by the US *CCDR* yields an Indirectly age-and-sex Standardized CoViD-19 Death Rate (*ISCDR*) for that population, with the US age-and-sex population distribution as the standard:

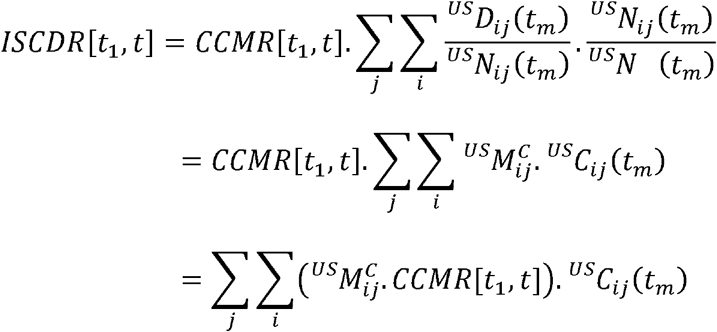

*CCMR* and *ISCDR* are again calculated for the period starting on the day of the first death in the population and ending on January 1^st^, 2021.

Last, life expectancy at birth provides a summary indicator of mortality in a population in a more intuitive metric (years) than these rates. A standard demographic technique allows to estimate the impact that *eliminating* a cause of death would have on life expectancy at birth.^11 12^ When a prior period life table (i.e., not factoring CoViD-19 mortality) is available, applying this technique backward allows to translate a cumulative CoViD-19-deaths forecast for the same period into a CoViD-19-induced *reduction* in male and female life expectancies at birth. The cumulative number of reported CoViD-19 deaths in 2020 were used to derive new male and female life expectancies at birth in 277 populations with extant life tables (155 countries, plus Italian regions, Spanish autonomous communities, Mexican and US states). Calculations required a previous projection of the male and female year-2020 life tables in these populations.

For countries, these were again derived from UN data, by interpolation between the 2015-20 estimates and 2020-25 projections. For sub-national populations, life tables available from national statistical institutes were extrapolated to 2020. Additional details on their calculation are described in the online supplementary materials of this article.

### Patient and Public Involvement

This research was done without patient involvement. Patients were not invited to comment on the study design and were not consulted to develop patient relevant outcomes or interpret the results. Patients were not invited to contribute to the writing or editing of this document for readability or accuracy.

## Results

To illustrate the properties of these indicators, we briefly describe results from the January 1^st^, 2021 updates of the CCSE and CDC data. (Full results for that day are available in the online supplementary materials of this article; updated results will continue to be uploaded to https://github.com/statsccpr/ind-cov-mort). For the period starting on the day of the first CoViD- 19 death observed in a population and ending on January 1^st^, 2021, the highest national values of the *CCDR* (given in deaths per thousand person-years) are found in five European nations (San Marino, 2.11; Belgium, 2.10; Slovenia, 1.63; Bosnia and Herzegovina, 1.57; North Macedonia, 1.54). Among the 20 nations with the highest values, only Peru (1.45), the USA (1.25), Mexico (1.24) and Argentina (1.16) are outside Europe. This list of nations, however, illustrates the issue of scale with small, densely populated nations exhibiting higher values than some larger nations, but possibly not than similarly-sized parts of these nations. If comparisons are based on subnational rather than national boundaries, *CCDR* values for populations of 5 million or more are higher for parts of Italy (Lombardy, 2.90), the USA (New Jersey, 2.65; New York, 2.39; Massachusetts, 2.27) and Spain (Madrid, 2.15) than for Belgium. Values for parts of Mexico (Mexico City, 2.06), Peru (Lima, 1.99), and Brazil (Rio de Janeiro, 1.85) and for five other US states (Illinois, Michigan, Pennsylvania, Arizona and Indiana) are also higher than for any nation besides Belgium (again among populations of 5 million or more).

The main motivation for the *CCDR* is not to compare CoViD-19 mortality across populations, however, but rather to compare CoViD-19 and overall mortality. Across the populations monitored here, the highest *CCDR* value to date for a period starting on the day of the first CoViD-19 death has been reached in New York (9.44 for the period ending on 4/25) where it exceeded the state’s most recent annual *CDR* (7.83 in 2017).^13^. The period *CCDR* remained above the 2017 *CDR* until May 20 (Figure 1). Ignoring competing risks between CoViD-19 and other-cause mortality, and seasonality and period trends in other-cause mortality, this indicates roughly equivalent mortality from CoViD-19 and from all other causes combined between March 14 (first death) and May 20.

**Figure 1:**
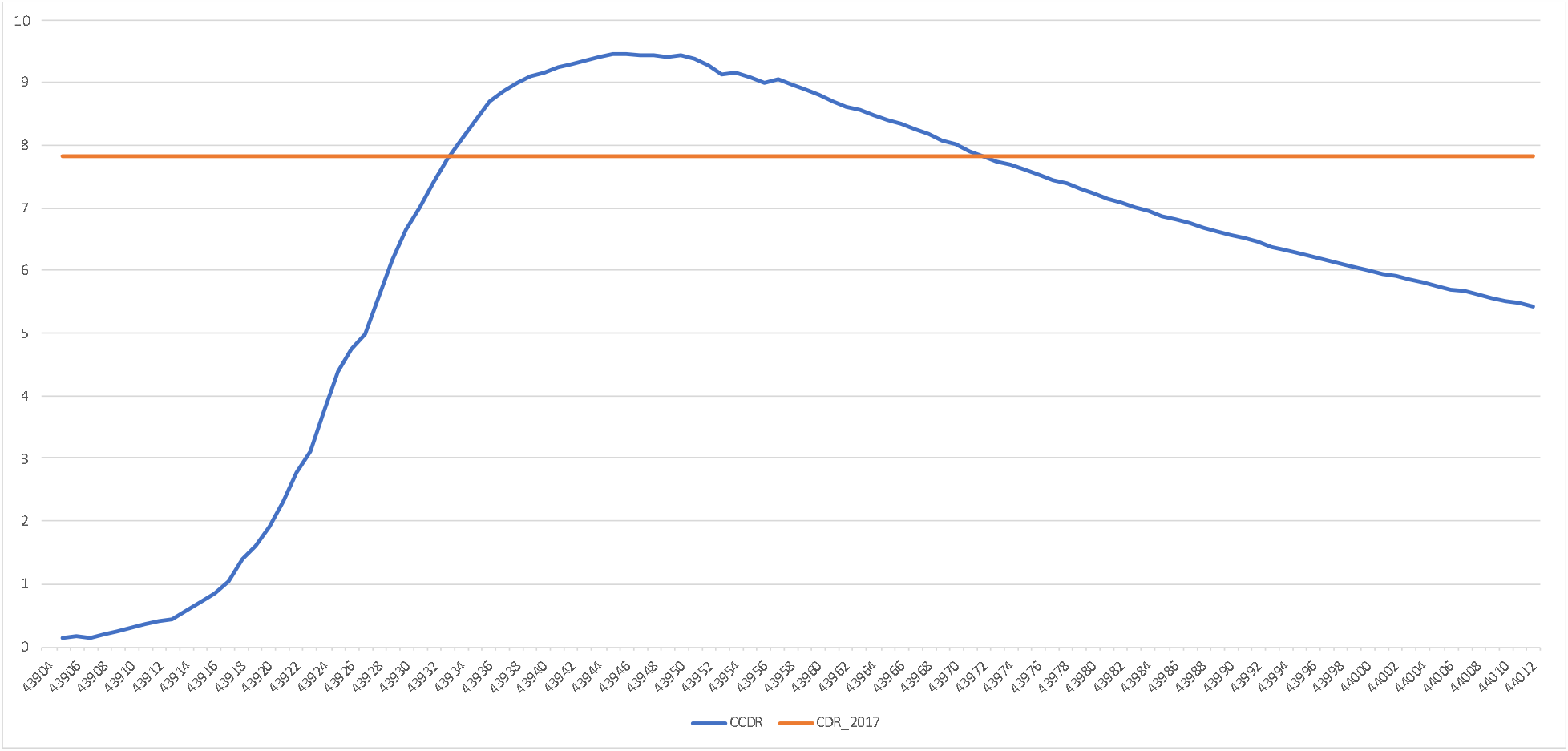
Estimated value of the period *CCDR*, New York (in deaths per 1,000 person-years, period starting on March 14 and ending on day shown on the horizontal axis) Sources: CDC (*CDR*) and authors’ calculations (*CCDR*, see technical appendix)

The effects of indirect age-standardization are illustrated in Figure 2, comparing current-period *CCDR* and *ISCDR* values for selected national and subnational populations (both in deaths per thousand person-years). By construction, the *CCMR* equals 1 and the *CCDR* and *ISCDR* are the same in the US, but the standardized *ISCDR* is lower than the unstandardized *CCDR* in Europe, whereas the standardized rate can be two to three times the unstandardized rate in Mexico and South American countries. The 20 highest values of the *ISCDR* are for eleven Mexican States and eight Peruvian *Departamentos*, ahead of Rio de Janeiro (Brazil). Among national and subnational units with a population size of 5 million or more shown in Figure 2, the highest value in Europe (Lombardy, 1.95) is lower than subnational values for Mexico (3.51) and four other Mexican states, Lima (Peru, 3.46), Rio de Janeiro (Brazil, 3.14) and four other Brazilian states, and New Jersey (2.52) and two other US states, as well as national values for Peru (2.68), Mexico (2.51), Bolivia (2.15) and Ecuador (2.05).

**Figure 2:**
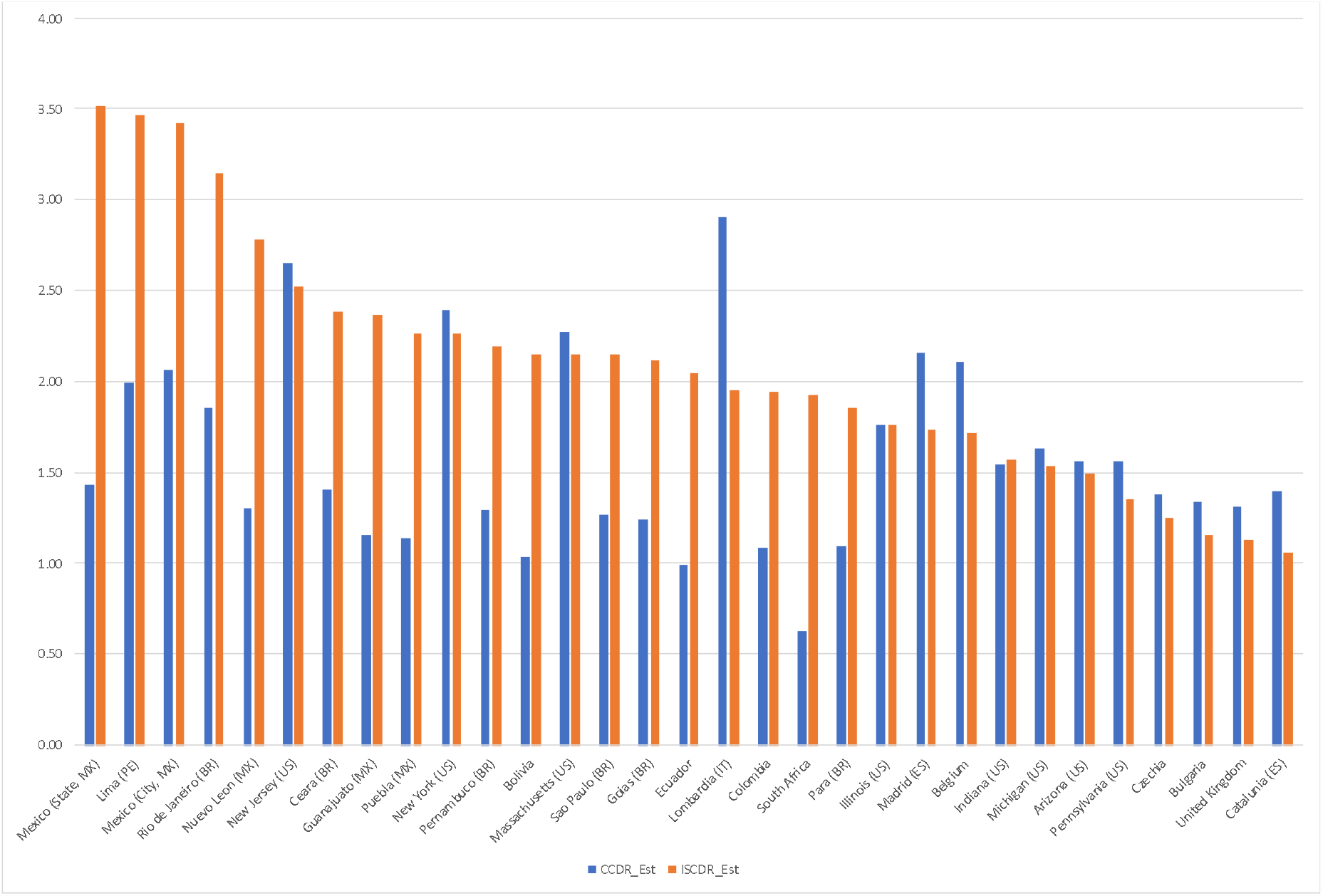
Estimated value of the *CCDR* and *ISCDR* (in deaths per 1,000 person-years), by national and subnational unit (20 *CCDR* and 20 largest *ISCDR* values for units with a population size of 5 million or more) Sources: Authors’ calculations (see technical appendix)

As for the mortality impact, reductions in 2020 life expectancies at birth of two years or more are estimated for two nations: Panama (2.22) and Peru (2.09). Subnational values were estimated within five nations and if reductions of two years or more were also estimated for Madrid (Spain), two Italian regions, two Peruvian *Departamentos*, and for four US states, values exceed two years for eleven Mexican states, foremost, Quintana Roo (includes Cancún, 3.93) and Baja California (includes Tijuana, 3.54, both values in years). Figure 3 shows reductions exceeding 1.3 years in 20 national and subnational units with a population size of 5 million or more.

**Figure 3:**
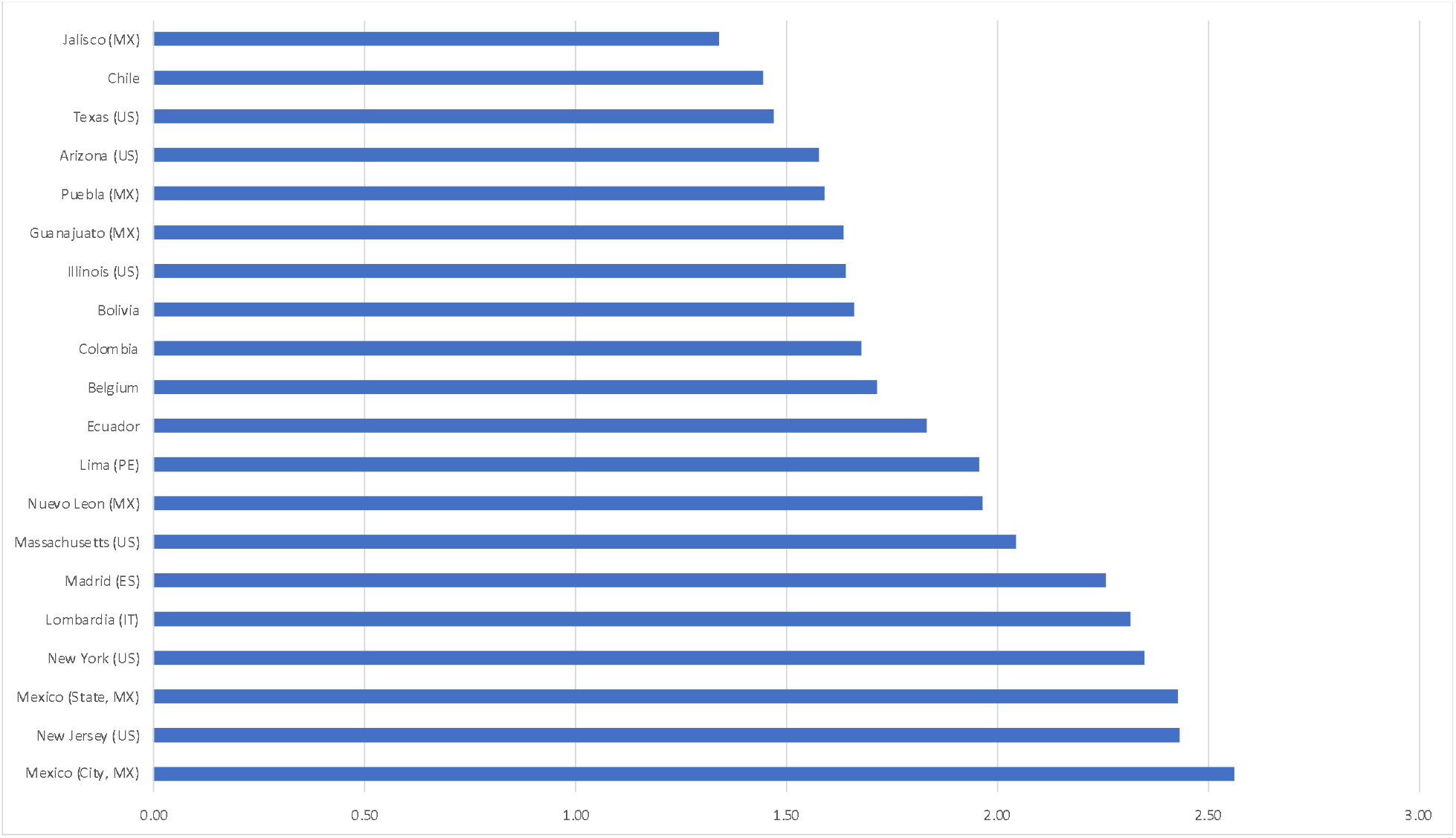
Estimated reduction in life expectancy at birth for year 2020, both sexes (in years), by national and subnational unit (20 largest reductions for units with a population size of 5 million or more) Sources: Authors’ calculations (see technical appendix)

Period life expectancy at birth is a summary indicator of mortality conditions across the lifespan that is available for all nations and each year since 1950 from the United Nations, and for earlier periods in a number of nations. This allows for comparing the mortality impact of CoViD-19 with prior reversals in the secular increase in life expectancies. An examination of the United Nations times series, for instance, suggests that next to the exceptional declines induced by mass homicides in Cambodia (1975-78) and Rwanda (1994), the largest annual declines in life expectancy at birth since 1950 took place in Eswatini (formerly Swaziland) during the worse years of the HIV pandemic (2.10 years between 1997 and 1998). Inducing declines in life expectancies for close to two decades in some countries, the HIV pandemic has had a much larger cumulative impact than CoViD-19 to date, but the fact that life expectancy at birth may decline by a larger amount in 2020 in a few national and subnational populations than in the most affected countries in any year during the HIV pandemic puts in perspective values shown in Figure 3. Moreover, while the estimated reduction for the USA (1.26) is lower than for the populations shown in Figure 3, the US life expectancy drop in 2020 will still be the largest since World War II, far exceeding declines in the worst year of the HIV epidemic (from 75.8 years in 1992 to 75.5 years in 1993), or the worst three years of the opioid-overdose crisis (from 78.9 years in 2014 to 78.6 years in 2017).^14^ As illustrated in Figure 4, CoViD-19 is estimated to reduce US life expectancy at birth in 2020 to its lowest level since 2005.

**Figure 4:**
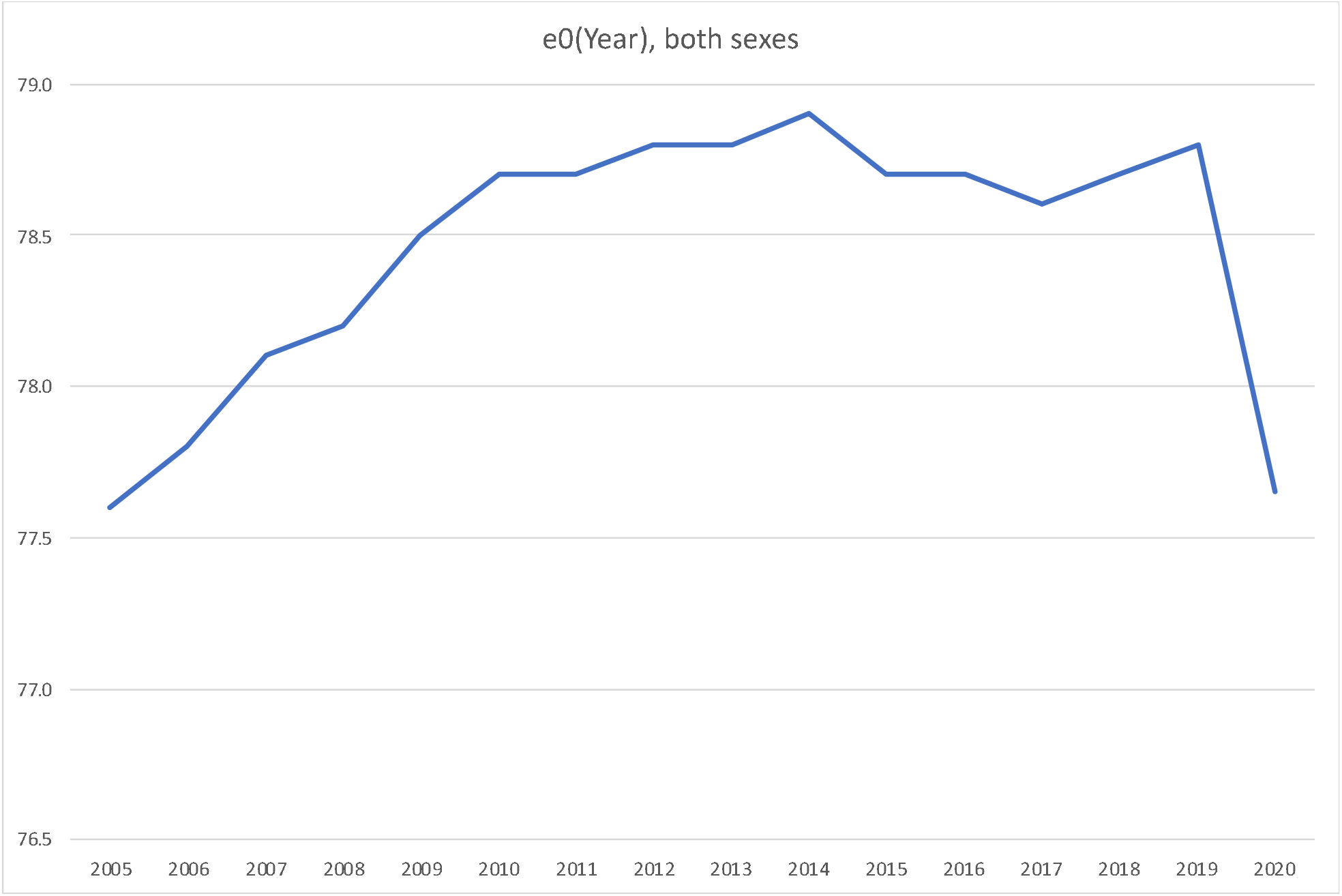
Estimated life expectancy at birth, US population, both sexes, by year Sources: CDC (2005-2018), UN and authors’ calculations (2019-2020, see technical appendix)

## Discussion

The results above illustrate the properties of different comparative indicators of CoViD-19 mortality. For comparisons across populations, the *ISCDR*, and *CCMR* on which it builds, control for three important factors that contribute to the cumulative count of CoViD-19 deaths in a population: the length of the period over which these deaths are cumulated, the size of the population, and its age-and-sex composition.

With respect to the first of these three factors, both the unstandardized and standardized rates are period indicators that increase and decrease as waves of the pandemic develop.

Contrary to the death per capita ratio, which can only increase over time, the period rates begin to decline when the daily number of additional deaths drops below its average for the period. This property of the period rates accurately reflects for CoViD-19 mortality a temporal dimension that can often be neglected for overall mortality. This also implies, however, that comparing *ISCDR* values across populations at too different durations of exposure to CoViD-19 would not be meaningful. As shown in Figure 1, this is more problematic early in the diffusion of the epidemic.

With respect to the second factor, comparing *ISCDR* values at the national or sub-national levels illustrates that dividing by population size does not make small and large populations fully comparable. National rates are but population-weighted averages of subnational rates. With person-to-person transmission and uneven population density, these rates can be expected to vary substantially across the territory of the largest countries, making it less likely that the national average will stand out in cross-national comparisons. While the national *ISCDR* is lower for Italy, Spain and the USA than for Belgium, each of these three nations has at least one subnational entity of roughly similar population size with a higher *ISCDR* than Belgium.

Disaggregation to smaller administrative units may allow for more meaningful comparisons, but might be impeded by data availability. In this respect, indirect standardization has the advantage of not requiring data on CoViD-19 deaths by age and sex that may not be available or reliable for smaller areas. As a reliable breakdown of CoViD-19 deaths *is* available from a number of European countries^15 16^ and US states, the *ISCDR* can actually be compared to a Directly age-and-sex Standardized CoViD-19 Death Rate (*DSCDR*) with the US age-and-sex population distribution as the standard. Comparing unstandardized with directly and indirectly standardized rate for the three European nations and the three US states with the highest *ISCDR* values, Figure 5 shows that the values of the indirectly standardized rate are typically very close to the corresponding values of the directly standardized rate.

**Figure 5:**
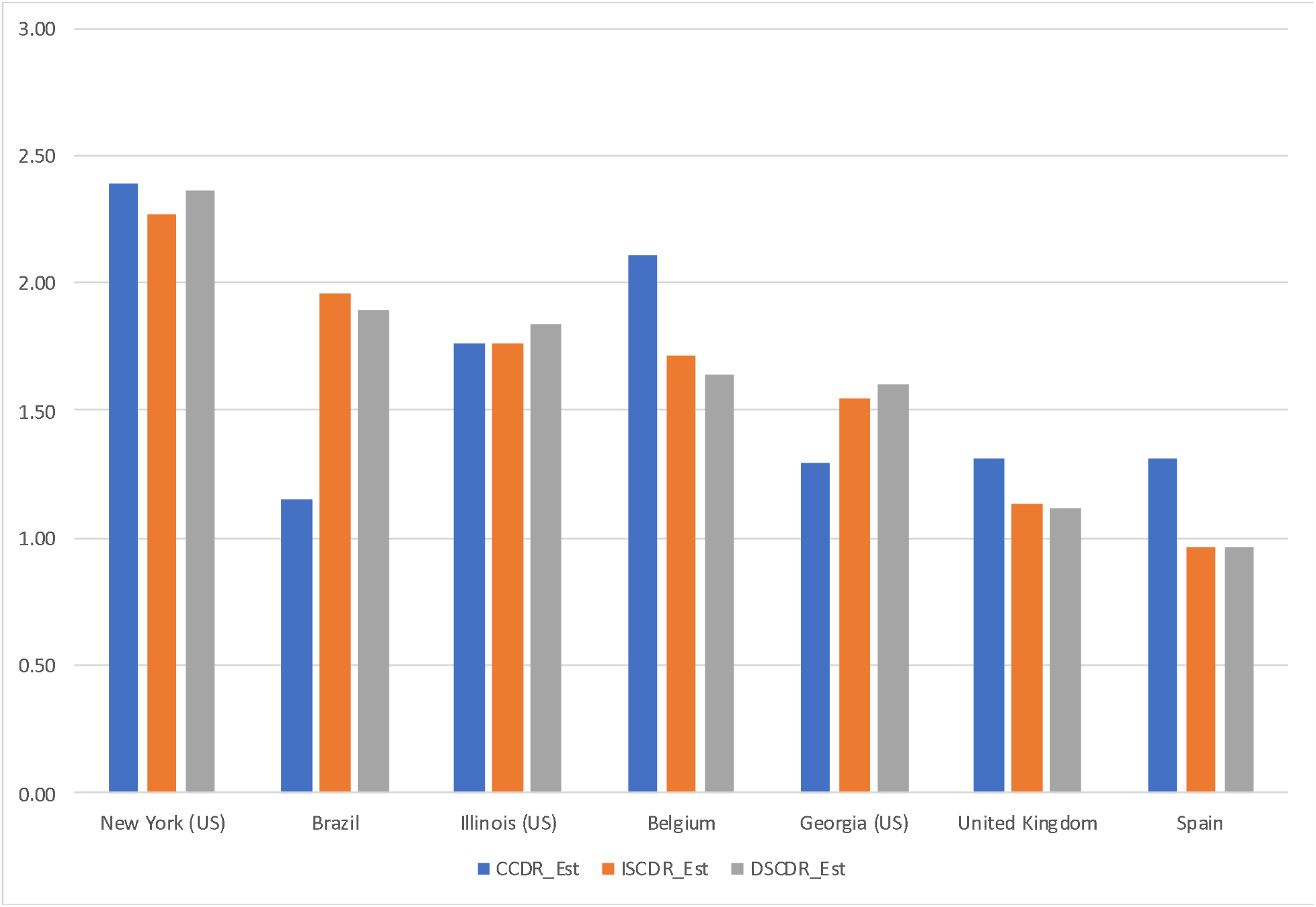
Estimated value of the *CCDR, ISCDR* and *DSCDR* (in deaths per 1,000 person-years), by selected nation and US state Sources: Ined, CDC, *Registro Civil* (Brazil)^17^ and authors’ calculations (see technical appendix)

Over time, sex- and age-specific rates of CoViD-19 mortality have become available for a larger and more diverse set of nations,^18^ providing a wider choice of possible standards. As is the case with the values of directly standardized rates, the values of indirectly standardized rates depend on the choice of a standard. While it is theoretically possible that the choice of a standard would also affect the rankings of directly or indirectly standardized rates, empirical regularities in mortality patterns across populations make this quite unlikely. The age patterns of CoViD-19 death rates available so far similarly exhibit remarkable regularities, with some modest variation in the slope of these age patterns at the oldest ages, probably due to the number of fatalities in nursing homes across Europe and the USA.^19^ Indirect standardization thus appears to provide a valid alternative to rank CoViD-19 mortality across populations when data limitations prevent direct standardization.

Variations in the slope of the age-specific rates of CoViD-19 mortality would also affect the estimated reductions in life expectancy at birth. If these rates increase less rapidly with age at the oldest ages in a population than they do in the USA, the age pattern of CoViD-19 deaths obtained here by multiplying the US age-specific rates and the population sizes of the different age groups would then be “older” than the actual age pattern. In turn, this would imply that the average number of years of life lost per CoViD-19 death and the total impact of CoViD-19 on life expectancy at birth is actually larger than estimated here. A simulation using the reported sex- and age-distribution of CoViD-19 for Brazil yielded a 1.67-year estimated reduction in life expectancy, however, only a 3% difference with the reduction estimated here (1.72 years, see Supplementary Information).

Another source of uncertainty originates in the role of preexisting conditions in CoViD- 19 mortality. While this role is well documented, data on CoViD-19 fatalities by preexisting conditions are even less commonly available than data CoViD-19 fatalities by age.^20^ One study suggests that the average number of years of life lost per CoViD-19 death might be overestimated by about 10% when preexisting conditions are not accounted for.^21^ This provides an order of magnitude for the upward bias that ignoring preexisting conditions might similarly induce in estimating reductions in life expectancies case here based solely on the age of CoViD- 19 victims.

Moreover, the illustrative results presented here make no adjustment for potential biases in the number of confirmed CoViD-19 deaths. Estimates of life-expectancy reductions based on these also assume no “indirect” effect of the pandemic on other-cause mortality. In populations with complete and timely registration of deaths, the reporting biases and indirect effects can be jointly assessed from the increase in overall mortality over past “benchmark” mortality levels. As noted in the introduction, CDC data continues to suggest that the cumulative number of CoViD- 19 deaths to date does not fully account for the overall increase in US mortality. However, the estimation of “excess” deaths directly or indirectly attributable to CoViD-19 can be quite sensitive to the choice of a benchmark period to represent past mortality conditions. Using the shiny app from the *Human Mortality Database*,^22^ the number of excess deaths for the first half of 2020 in France, for instance, can be estimated to reach approximately 40,000 when years 2000 through 2019 are used to estimate benchmark mortality, whereas using only the most recent year, 2019, for that mortality benchmark, the estimate drops below 30,000—the number of confirmed CoViD-19 deaths over the same period.^23^

The results presented here to illustrate the properties of these period indicators can easily be customized for different periods, different geographical scales, or to assess their robustness to these different sources of uncertainty. For tracking the pandemic, for instance, estimating *CCDR* and *ISCDR* values for more recent periods than the period starting with the first CoViD-19 death and at a smaller scale than the first subnational division would be necessary. While this can be done with life-expectancy reductions as well, the value of life expectancy for a short period in a small geographical area becomes difficult to interpret and additional measures might become better suited to express the effect of CoViD-19 on longevity.^24 25 26^ The *ISCDR* and life-expectancy reductions are the least data-demanding of the summary indicators of mortality conditions that allow for comparisons across populations, however, and as crude death rates and life expectancy estimates are widely available, the *CCDR* and life-expectancy reductions readily allow for temporal comparisons with other-cause mortality.

## Supporting information

Technical Appendix

Full Results

## Data Availability

All data are available online and can be accessed directly or through a web scraping routine that we provide at https://github.com/statsccpr/ind-cov-mort

https://www.arcgis.com/apps/opsdashboard/index.html#/bda7594740fd40299423467b48e9ecf6

https://covid19.healthdata.org/

https://www.cdc.gov/nchs/nvss/vsrr/COVID19/index.htm

## Acknowledgment

The authors benefited from facilities and resources provided by the California Center for Population Research at UCLA (CCPR), which receives core support (P2C-HD041022) from the Eunice Kennedy Shriver National Institute of Child Health and Human Development (NICHD). The authors thank Patrick Gerland for clarifications regarding UN demographic data, Hiram Beltrán-Sánchez for additional data for Mexico, and Enrique Acosta, Lars Ängquist, Sam Preston, Jason Kerwin and Piedad Urdinola for comments on an earlier draft.

## Data Sharing Statement

Additional data are available on the Github repository: https://github.com/statsccpr/ind-cov-mort

## Ethics

This study has no human subjects. Analyses are based solely on publicly available online data on anonymous, deceased individuals.

## Funding

The authors benefited from facilities and resources provided by the California Center for Population Research at UCLA (CCPR), which receives core support (P2C-HD041022) from the Eunice Kennedy Shriver National Institute of Child Health and Human Development (NICHD).

## Notes

### Competing Interest Statement

The authors have declared no competing interest.

### Author Declarations

Publicly available data only

### Summary of Updates

This revised version uses updated JHU estimates for January 1, 2021

