## Supplementary material for "Beyond Deaths per Capita: Comparative CoViD-19 Mortality Indicators": Technical Appendix

**Technical Appendix**

*Updated Version, 1/5/2021*

### Part A. Mortality Indicators

#### Section 1. Period Crude CoViD-19 Death Rate (*CCDR*)

1.1 Get current estimate date and cumulative number of CoViD-19 deaths by UN country/territory, , and subnational entities with at least one reported case by June 1, 2020 in Brazil, China, Italy, Mexico, Peru, Spain and the USA (all locations thereafter) from: https://coronavirus.jhu.edu/

1.2 Get date of first CoViD-19 death and total mid-2020 population size for all locations in (1.1) (see part B for example)

1.3 Calculate exposure in person-years for all locations in (1.1) as:

$$N.T$$

where *N* is total population size in (1.3) and *T* is year-to-date duration in year converted from dates in (1.1) & (1.2)

1.4 Calculate the estimated period Crude CoViD-19 Death Rate (*CCDR*) for all locations in (1.1) and projected *CCDR* for all locations in (1.2) as ratios of deaths in (1.1) & (1.2) to exposure in (1.3)

#### Section 2. Comparative CoViD-19 Mortality Ratio (*CCMR*)

2.1 Get report date and number of reported CoViD-19 deaths by sex and age group from: https://data.cdc.gov/NCHS/Provisional-COVID-19-Death-Counts-by-Sex-Age-and-S/9bhg-hcku

2.2 Get the mid-2020 population size by age groups, ${}_nN_x$, for each sex and the same age groups as in (2.1) for all locations in (1.1) (see part B for example)

2.3 Calculate age-and-sex-specific CoViD-19 death rates for the USA using the estimate date of first CoViD-19 death and estimated number of CoViD-19 deaths in the country in (1.1), the distribution of deaths by sex and age group in (2.1) and the mid-2020 population by sex and age group in the country in (2.2) as:

$${}_nD_x^C \,/\, {}_nN_x.T$$

where (separately for males and females) ${}_nN_x$ is the mid-2020 population in age group *x* to *x*+*n* in (2.2), *T* is the duration of exposure in (1.4) and ${}_nD_x^C$ is the number of CoViD-19 deaths in (1.1) multiplied by the ratio of deaths in the age group to total deaths in (2.1)

2.4 Calculate estimated counterfactual numbers of CoViD-19 deaths for all locations in (1.1) using the sex and age-specific CoViD-19 death rates for the USA in (2.3) and the mid-2020 population by sex and age group in (2.2)

2.5 Calculate the Comparative CoViD-19 Mortality Ratio (CCMR) for estimated numbers of CoViD-19 deaths for all locations in (1.1) as the ratio of the actual estimate in (1.1) to the corresponding counterfactual number in (2.4)

#### Section 3. Estimated Reduction in 2020-Life Expectancies

3.1 Get period life-table age-specific death rates (${}_nm_x$) and survival probabilities (${}_np_x$) for year-2020 for each country in (1.1) (see part B for example)

3.2 Calculate the age-specific ratio of updated to previously projected deaths from all causes in 2020 for each country in (1.1) as:

$$ {}_nR_x = \frac{{}_nm_x . \left( {}_nN_x - \left( (1 - \overline{t_m}) . {}_nD_x^C \right) \right) + {}_nD_x^C}{{}_nm_x . {}_nN_x} $$

where ${}_nm_x$ is the age-specific death rate in the previously projected year-2020 life table from (3.1), ${}_nN_x$ is the mid-2020 population by age group from (2.2), ${}_nD_x^C$ is the estimated number of CoViD-19 deaths in the age group obtained by multiplying the total for the Country in (1.2) by the ratio of deaths in the age group to total deaths in the USA in (2.1) and $\overline{t_m}$ is the fraction of a year corresponding to the average time of CoViD-19 deaths estimated here as the mid-point between the first CoViD-19 death in the country and the end date of the projection (January 1, 2021 as of this writing)

${}_nD_x^C$ is obtained by multiplying ${}_nN_x$, first, by the corresponding age-and-sex-specific CoViD-19 death rate for the USA in (2.3) and, second, by a scaling factor, identical across all sex- and age-groups, to bring the sum of the estimated number of CoViD-19 deaths across all sex- and age-groups to equal the total estimated number of CoViD-19 deaths in the population. (The scaling factor is the ratio of the total estimated number of CoViD-19 deaths in the population divided by the sum of the products of ${}_nN_x$ by the corresponding age-and-sex-specific CoViD-19 death rate for the USA)

3.3 Calculate age-specific survival probabilities in the new projected year-2020 life table for each country in (1.1) from (3.1) & (3.2) using Chiang (1968) formula:

$$ {}_n\overset{*}{p}_x = {}_np_x^{\;{}_nR_x} $$

3.4 Calculate the age-specific number of years lived after age *x* for individuals dying in the age interval (${}_na_x$ values) in the new projected year-2020 life table for each country in (1.1) from its corresponding value in the previously projected year-2020 life table derived from (3.1) and the life table relationship:

$$ {}_na_x = \frac{1}{{}_nm_x} - n.\frac{{}_np_x}{1 - {}_np_x} $$

and from (3.2) & (3.3) using the Preston et al. (2001: 84) formula:

$$ {}_n^*a_x = n + \left( {}_nR_x . \frac{{}_nq_x}{{}_n^*q_x} . \left( {}_na_x - n \right) \right) $$

where ${}_nq_x$ is 1-${}_np_x$, and

$$ {}^*a_{85+} = \frac{a_{85+}}{R_{85+}} $$

3.5 Calculate new values of life expectancies ($e_x^o$ values) in the year-2020 life table for all locations in (1.2) starting with $e_x^o$=*$a_{85+}$ in (3.4) and then using values in (3.3) & (3.4) with the life table relationship:

$$ e_x^o = {}_np_x . \left( e_{x+n}^o + n \right) + {}_na_x . \left( 1 - {}_np_x \right) $$

3.6 Calculate the difference between the new values of life expectancies in year-2020 life table in (3.5) and the original values derived from values in (3.1) for all locations in (1.2) and the life table relationship:

$$ e_x^o = \left( {}_np_x . e_{x+n}^o \right) + \frac{1}{{}_nm_x} . \left( 1 - {}_np_x \right) $$

#### Part B. Demographic Parameters

Section 1. Mid-2020 Population Size

1.1 (Step 1.3 in part A) Total mid-2020 population size for each UN country and territory was obtained from the “Total Population” file at:
https://population.un.org/wpp/Download/Standard/CSV/
1.1.a For provinces in China, the mid-2020 population size for the country was multiplied by the ratio of the 2019-year-end total population estimates for the province divided by the corresponding estimates for the country obtained at:
http://data.stats.gov.cn
1.1.b For US states, the total population size was obtained by adding across sex- and age-groups, with the mid-2020 size of each sex- and age-group for the country being multiplied by the ratio of the 2018-sex-and-age-group sizes estimated for the state divided by the corresponding estimate for the country obtained at:
https://data.census.gov/cedsci/table?q=United%20States&g=0100000US&tid=ACSDP1Y2018.DP05&hidePreview=true&table=DP05
1.1.c For Brazilian states, the mid-2020 population size for the country was multiplied by the ratio of the 2019-year-end total population estimates for the state divided by the corresponding estimates for the country obtained from the Brazil Statistical Office (IBGE) at:
https://www.ibge.gov.br/en/cities-and-states.html?view=municipio
1.1.d For Italian regions, the total population size was obtained by adding across sex- and age-groups, with the mid-2020 size of each sex- and age-group for the country being multiplied by the ratio of the 2019-age-group sizes estimated for the region divided by the corresponding estimate for the country obtained from IStat at:
http://demo.istat.it/tvm2016/index.php

1.1.e For Spanish autonomous communities, the total population size was obtained by adding across sex- and age-groups, with the mid-2020 size of each sex- and age-group being multiplied by the ratio of the 2019-age-group sizes estimated for the community divided by the corresponding estimate for the country obtained from Instituto Nacional Estadistica (INE) at:
https://www.ine.es/dyngs/INEbase/en/categoria.htm?c=Estadistica_P&cid=125473471098
4
1.1.f For Mexican states, the total population size was obtained by adding across sex- and age-groups, with the mid-2020 size of each sex- and age-group being multiplied by the ratio of the 2010-age-group sizes estimated for the state divided by the corresponding estimate for the country obtained from Instituto Nacional de Estadistica y Geografía (INEGI) at:
https://en.www.inegi.org.mx/temas/estructura/default.html#Tabulados
1.1.g For Peruvian *departamentos*, the total population size was obtained by adding across sex- and age-groups, with the mid-2020 size of each sex- and age-group being multiplied by the ratio of the 2017-age-group sizes estimated for the state divided by the corresponding estimate for the country obtained from El Instituto Nacional de Estadísticañ e Informática (INEI):
https://www.inei.gob.pe

1.2 (Step 1.2 in part A) Dates of first CoViD-19 case and death for UN countries and territories and for provinces in China were retrieved from the World Health Organization's daily situation reports at https://www.who.int/emergencies/diseases/novel-coronavirus-2019/situation-reports/
1.2.a. For other sub-national populations, dates were obtained from the Institute for Health Metrics and Evaluation at https://covid19.healthdata.org
1.2.b. For Peruvian *departamentos*, dates were not available from IHME and assumed to all be the same as the date of the first CoViD-19 death in the country.

1.3 (step 2.2. in part A) For UN countries and territories with a population size over 90,000,mid-2020 population size by sex- and age-group was obtained from the "Population by Age and Sex" file at: https://population.un.org/wpp/Download/Standard/CSV/
with the following adjustments:
1.3.a Number of infants (under age 1) for each of these UN countries and territories was obtained from the "Annual Population by Age – Both Sexes" file at:
https://population.un.org/wpp/Download/Standard/CSV/
1.3.b Number in age group 1-4 for each of these UN countries and territories was obtained as the difference between the number in the first age group (age 0-4) in the population by 5-year age groups in (1.3) above and the number of infants in (1.3.a) above
1.3.c Numbers in age groups 5-14 to 75-84 for each of these UN countries and territories were obtained by adding the numbers in two consecutive age groups (e.g., ages 5-9 and ages 10-14) in the population by 5-year age groups in (1.3) above
1.3.d Number in age group 85 and over for each of these UN countries and territories was obtained by adding the numbers in the last four age groups (i.e., ages 85-89, 90-94, 95-99 and 100+) in the population by 5-year age groups in (1.3) above

1.3.e For sub-national populations in Brazil and China, the total population size from (1.1.a) or (1.1.c) above was multiplied by the ratio of the national mid-2020 estimates for the sex- and age-group and for the total population (for the USA, Italy, Spain, Mexico and Peru, see (1.1.b), (1.1.d), (1.1.e), (1.1.f) and (1.1.g) above)

Section 2. Calendar-Year-2020 Period Life Table Values

2.1 (Step 3.1 in part A) The period life-table age-specific survival probabilities (${}_np_x$) for year-2020 for each country in (1.3) above are obtained from the corresponding values in the estimated 2015-20 and projected 2020-25 life tables in the "Life table survivors (lx) at exact age x - Both Sexes" file at: https://population.un.org/wpp/Download/Standard/CSV/

2.1.a Age-specific survival probabilities (${}_np_x$) in the estimated 2015-20 and projected 2020-25 life tables for each country in (1.3) above are obtained from the number of survivors by age ($l_x$) and the life table relationship:

$$ {}_np_x = \frac{l_{x+n}}{l_x} $$

2.1.b Period life-table age-specific survival probabilities (${}_np_x$) for year-2020 for each country in (1.3) above are obtained as:

$$ {}_np_x[2020] = \sqrt[2]{{}_np_x[2015-2020].\ {}_np_x[2020-2025]} $$

2.2 (step 3.1 in part A) The period life-table age-specific death rates (${}_nm_x$) for year-2020 for each country in (1.3) above are obtained from the corresponding values in the estimated 2015-20 and projected 2020-25 life tables in the "Life table survivors l(x) at exact age x" and "life expectancy at exact age x" files at: https://population.un.org/wpp/Download/Standard/CSV/

2.2.a Age-specific death rates (${}_nm_x$) in the estimated 2015-20 and projected 2020-25 life tables for each country in (1.3) above are obtained the male/female number of survivors by age ($l_x$) and the male/female life expectancy by age ($e_x^o$) and the life table relationships:

$$ {}_nm_x = \frac{l_x - l_{x+n}}{(l_x . e_x^o) - (l_{x+n} . e_{x+n}^o)} $$

and

$$ m_{x+} = \frac{1}{e_x^o} $$

2.2.b Period life-table male/female age-specific death rates (${}_nm_x$) and survival probabilities (${}_np_x$) for year-2020 for each country in (1.3) above are obtained as:

${}_nm_x[2020] = ({}_nm_x[2015\text{-}20] + {}_nm_x[2020\text{-}25]) /2$

2.2.c For US states, age-specific death rates (${}_nm_x$) for 2016 at ages above age 25 were obtained from: https://wonder.cdc.gov/controller/datarequest/D140. Under age 25, age-specific death rates are unreliable in some states and the rates in all states were thus assumed to equal the rate at the same age in the country (the assumption should have negligible impact on CoViD-19 mortality assessments). The 2016 age-specific death rates for each sex, age group and state were thus prorated using the ratio of 2020/2016 age-specific death rate for the same sex and age group in the country.

2.2.d For Italian regions and Spanish autonomous communities, 2018 (Italy) and 2019 (Spain) life tables were obtained from IStat and INE (see 1.1 above). The 2018 (or 2019) age-specific death rates for each sex, age group and region/community were prorated using the ratio of 2020/2018 (or 2019) age-specific death rate for the same sex and age group in the country. The 2020 life tables were then completed assuming no change in the ${}_na_x$ from 2018 (or 2019)

2.2.e For Mexican states, 2020 life tables were taken directly from those produced by Partida Bush & García Guerrero (2018) for the UN Population Fund:

Partida Bush, V.G. and García Guerrero, V. (2018). Proyecciones de la Población de México y sus Entidades Federativas 2016–2050. Consejo Nacional de Población, Fondo de Población de Naciones Unidas.

2.2.f For Peruvian *departamentos*, 2020 life tables were interpolated as described in 2.2.a & 2.2.b from projected life tables for 2015-2020 and 2020-2025 obtained from El Instituto Nacional de Estadísticañ e Informática (INEI) at:

http://proyectos.inei.gob.pe/web/biblioineipub/bancopub/Est/Lib0901/index.htm
